# Quality of Life in Parkinson’s Disease: Insights from a Single-Session Focus Group in Southwestern Ontario

**DOI:** 10.1101/2025.04.04.25325279

**Authors:** Daniela Carvalho, Devorah Dolman, Julia Kaf-Alghazal, Hafsa Mir, Faraj Haddad, Hossein Noyan

## Abstract

**Background:** Parkinson’s Disease (PD) is a chronic illness that profoundly impacts quality of life (QoL). While many qualitative studies on QoL in PD have been conducted in different countries and cities around the world, the impact on QoL varies by region and it is important to explore the diverse factors influencing these differences.

**Objectives:** This pilot study aims to explore the impact of PD on QoL for patients in Southwestern Ontario, Canada. These individuals may encounter unique challenges not currently addressed by the literature, potentially affecting their overall QoL.

**Methods:** A single-session focus group was conducted in a rural town in Southwestern Ontario, Canada. Line-by-line reflexive coding was used to identify iterative themes.

**Results:** Seven themes were explored from the participant’s contributions, with key themes focusing on patients’ experiences navigating the healthcare system, the impact of non-motor and motor symptoms, and the role of social support in their QoL.

**Conclusion:** Living with PD presents a variety of unique challenges that must be considered throughout future research and policy implementation. This pilot focus group study provides an in depth discussion of the key challenges faced by individuals with PD In Southwestern Ontario. The study is limited in reliability and generalizability due to the small sample size and homogeneity of participants. The initial exploratory findings can be used as a foundation for future research to expand on key themes with additional focus group sessions or complementary methodologies.

## Introduction

Parkinson’s Disease (PD) is a prevalent neurodegenerative disorder affecting over more than 100,000 Canadians.^1^ PD is characterized by motor and non-motor symptoms (NMS) that worsen over time, significantly impacting patients’ quality of life (QoL).^2–4^ QoL is a subjective evaluation of overall life satisfaction and well-being, reflecting an individual’s perceptions of their life within the context of their goals, expectations, and concerns.^5–7^ QoL encompasses a wide range of domains, and it is crucial in chronic illness management as it reflects individuals’ subjective physical, mental, emotional, and social well-being.^8^ Understanding QoL in PD is essential as it reflects the broader impact of living with a chronic condition.

Standardized tools like the 39-item Parkinson’s Disease Questionnaire (PDQ-39) and the 8-item Parkinson’s Disease Questionnaire (PDQ-8) are commonly used to evaluate QoL in PD.^9,10^ These instruments measure perceived QoL across eight domains: mobility, daily living activities, emotional well-being, social support, stigma, cognition, communication, and bodily discomfort.^9^ However, these validated and structured tools may not fully capture the complexity of PD patients’ experiences.^11^ The rigid nature of these tools can limit the depth of responses, potentially overlooking significant aspects of QoL that are not easily quantifiable. Qualitative research methods, such as focus groups, allow for a deeper exploration of these dimensions, enabling patients to express their subjective experiences more freely.^12^

Focus group studies are valuable for understanding the perspectives of individuals with chronic illnesses, such as PD.^2,13^ These discussions enable patients to share their experiences in a supportive environment, providing insights into specific aspects of how their QoL is affected.^14^ Several qualitative studies have revealed themes consistent with the domains assessed by the PDQ-39 and PDQ-8.^2,11,12,14^ However, they have also uncovered additional themes often overlooked by quantitative measures, such as environmental factors like access to healthcare resources and geographic location.^2, 15, 16^ For instance, a study by Rosqvist et al. (2021) found that PD patients with access to specialized care, such as neurologists or PD nurses, reported higher QoL, highlighting the importance of accessible specialized care.^15^

While numerous studies have explored QoL in PD patients using standardized surveys and focus groups, most qualitative studies have been conducted in the United States (U.S.), which limits the applicability of these findings to other countries with different healthcare systems, including Canada, where the current study was conducted. Furthermore, the limited Canadian studies primarily focus on urban centers, neglecting the experiences of PD patients in smaller cities and rural regions. In Ontario, the most populous province in Canada, over two million residents lack a general practitioner (GP), forcing reliance on walk-in clinics or community health centers for primary care.^19^ Given that Ontario faces significant physician shortages and long wait times for specialized care, patients in non-metropolitan areas may experience a unique barrier that impact their QoL. ^2, 12, 15, 17, 18^ PD patients often wait at least one year to see a neurologist, and lacking a GP can further delay diagnosis and treatment, exacerbating symptoms and negatively impacting QoL.^20, 21^ These challenges are compounded for individuals in rural and remote regions, where access to specialized care is limited.^22^ Despite this knowledge, there is minimal research addressing these regional disparities. This study aims to bridge this gap by conducting a focus group in Woodstock, Ontario, to explore the lived experiences of PD patients and identify region-specific challenges in accessing care and managing their condition. Woodstock is a small city in Southwestern Ontario with a population of approximately 50,000 as of 2024 and has a regional hospital but lacks specialized care.^23^ The primary aim of this study is to generate insights into how PD impacts the QoL for people in Ontario, Canada. Currently, there is very limited research focused on investigating the unique challenges and barriers faced by Canadians living with PD in this region. This creates a significant gap in our understanding of the factors affecting the QoL for the PD population. Understanding how QoL is impacted across various domains is an important step to addressing these issues in disease management and policy implementation.

## Methods

### Research Design

This exploratory qualitative study utilized direct engagement via a focus group consisting of individuals with PD. Since QoL reflects perceptions regarding one’s own position in life within the social context in which they live, it is important to understand the perspectives of the target population and gather information directly from them.^24^ A focus group is the suitable methodology to meet this objective. Historically, studies investigating QoL within specific groups of interest have employed the use of focus groups in lieu of other methodologies such as surveys as a means of focusing on lived experience and encouraging more in-depth discussions about the unique needs of the target population.^24^ Additionally, we determined that direct engagement via a focus group was the most amenable way to collect data from geriatric populations, where filling out surveys or navigating online technology could become too burdensome. Additionally, the focus group was mutually beneficial because it acted as a support session whereby participants supported one another.

A single-session focus group was conducted to gather preliminary insights into the challenges faced by individuals with PD in Southwestern Ontario. This approach allowed for the initial exploration of key themes, informing potential directions for future, more extensive studies. The goal was to better understand participant experiences, perspectives, feelings, concerns, and other factors that impact their daily living.^25, 26^ By fostering direct and in-depth discussions, the focus group can illuminate the unique voice of the target population.

The questions used to assess QoL and guide the discussions of this focus group were based on the Parkinson’s Disease Questionnaire (PDQ-8) and covered the eight domains of QoL as outlined by the validated questionnaire. There were 8 questions that were discussed during the focus group. Each question was informed by the PDQ-8 domains. However, each question was tailored to become more open-ended and more conducive to generating discussion amongst the study participants. These questions were modified to capture unique, context specific experiences from our sample and suit the needs of the Ontario population. The PDQ-8 was used to guide the questions of this study group as psychometric evaluations of the instrument have been shown to be robust and reliable cross culturally, and in many diverse populations, including Canadian samples.^27–29^ By using the PDQ-8 to guide the topics discussed in the study focus group, we will ensure that the data collected can be accurately coded to answer the outlined research question of this study.

### Ethical Considerations

The study was reviewed and approved by the University of Western Ontario’s Health Science Research Ethics Board (Project ID:124356). Written informed consent was obtained from all participants before their involvement. Participants were informed they could withdraw at any time without facing any consequences. Confidentiality of the data and personal information collected was ensured, with all identifiable documents only accessible by the principal investigator and the research team.

### Participants

Participants were recruited through the Parkinson’s Society of Southwestern Ontario (PSSO), a local non-profit organization that supports individuals living with PD in Southwestern Ontario, including monthly support groups. Support groups allow people coping with the psychosocial aspects of PD to give and receive support. For this study, members from the Woodstock, Ontario support group and their care partners were invited to participate. Invitations were sent via email by the PSSO’s regional community engagement coordinator, along with a study advertisement detailing the focus group session. This approach allowed participants to share their experiences in a familiar setting similar to their regular monthly support meetings.

Eligible individuals were required to be members of PSSO, have a formal PD diagnosis, reside in Southwestern Ontario, and be fluent in English. Participants were excluded if they were unable to be physically present for the focus group session. Caregivers could participate as informants but were limited to reflecting the participant’s perspective. Any contributions regarding the caregivers’ own experiences were excluded from the analysis for the purpose of this study, as the experience of patients with PD was the main objective.

The study aimed to recruit eight to twelve participants, aligning with the optimal size for traditional focus groups.^2, 30^ Seven PD participants expressed interest, and six attended, along with three caregivers. Among the six participants, five identified as male and one as female, with ages ranging from 60 to 80 years. All participants identified as Caucasian or having European ancestry, with three holding a college or university degree and the remaining three having completed high school. All participants were retired and married. Time since diagnosis ranged from two to ten years.

### Data Collection

The focus group session was held at the PSSO facility in Woodstock, a centrally located town in Oxford County, Southwestern Ontario, with a population of approximately 50, 000 people.^31^ The facility was chosen for its accessibility, comfort, and familiarity, as it is the regular meeting place for monthly support group sessions. All participants were residents of Woodstock. The session lasted approximately 90 minutes with a 15-minute break halfway through.

Four moderators facilitated the session. Two led the discussion by pre-determined questions, while the other two transcribed participants’ responses verbatim. At the start of the session, moderators reviewed the Letter of Information and Consent and obtained written and verbal consent from all participants. Caregivers were reminded that their contributions should reflect the perspective of the PD patient they accompanied.

Following consent, participants completed a Demographics Questionnaire to gather basic sociodemographic information such as age, gender, sex, race, ethnicity, time since PD diagnosis, employment status, marital status, and educational background. This provided context for understanding the sample population. An audio recording of the session was made using the Zoom Corporation H4n Handy Recorder to ensure that all shared information was accurately captured for further analysis. Moderators posed questions to explore participants’ experiences with PD, focusing on several QoL domains identified in the literature. The session concluded with a debrief, allowing participant to share any final thoughts or questions. The Demographic Questionnaire and the structured questions are available upon request.

## Data Analysis

Thematic analysis followed the six steps outlined by Braun & Clarke (2006).^32^ This study did not aim to achieve full data saturation, as the focus was on depth of insight rather than exhaustive thematic coverage. The initial phase of the thematic analyses per these guidelines involved transcribing the audio recording verbatim using NVivo (QSR International, version 14.23.0, March 2023). The NVivo transcript was exported into a Microsoft Word document, to enable multiple researchers to collaborate simultaneously. The next phase involved selection of keywords, where the researchers highlighted and identified recurring sentences, patterns or terms to encapsulate the perceptions of the participants directly from the transcript on the Word document. The third step involved coding of the transcript document, which was evenly distributed among four researchers (DD, DC, JK, HM). A reflexive coding method was adopted where codes are not fixed at the start but develop dynamically as the researchers deepen their understanding of the data. The codes were then categorized into themes by two of these researchers (DC, HM), who alternated sections to avoid analyzing their own assigned portions from the coding phase to mitigate researcher biases and account for inter-coder reliability. In the thematic analysis and conceptualization phases, patterns in the data and quotes were organized into distinct, non-overlapping themes and sub-themes using a table in Microsoft Excel. Final themes were individually validated by all researchers to achieve consensus. This table ultimately became the codebook for this study. Any discrepancies in codes or themes were resolved through discussion among the research team, contributing to the finalized codebook. A reflexive framework was used, allowing themes to emerge directly from the transcript. This process led to a unique conceptual model specific to the experiences of individuals with PD.

## Results

One focus group session was held, involving six participants with PD. The average age of the six participants was 73.5 years (SD 8.12) and time since diagnosis was 4.8 years (SD 3.06). Five of the six participants were male. The focus group session revealed insights regarding experience of individuals living with PD, from which 7 general themes were identified: (1) mobility issues; (2) impact on activities of daily living; (3) emotional wellbeing; (4) stigma; (5) social support; (6) other related symptoms and (7) healthcare navigation.

## Discussion

The preliminary group session yielded insights into the lived experiences of individuals with PD, contributing to the result of our study. Participants shared personal challenges, adaptations, perspectives, and perspectives on navigating daily life with PD, shedding light on both the emotional and physical aspects of their journey with the disease. These firsthand accounts not only deepen our understanding of the disease’s impact on individuals in Woodstock, Ontario, but highlight a need for further research and investigation for more widespread patient populations to target potential areas for intervention. The following section explores themes that emerged from the discussion, offering a comprehensive view of the challenges faced by these individuals with PD.

### Mobility

The findings from the pilot focus group study highlight the impact of PD on mobility and daily function, particularly regarding progressive physical challenges such as stiffness, tremors, reduced dexterity, and handwriting difficulties. Participants described a range of adaptive strategies, including time management, gradual movements, and repetitive exercises, to mitigate these effects. However, despite the efforts, certain tasks remain difficult or even impossible, leading to a perceived loss of independence in some areas. For instance, one participant emphasized the need for a slow, deliberate approach to movement, particularly in tasks like getting out of bed, to avoid mobility limitations. Others noted that fine motor impairments, such as cramping and decreased dexterity, require dedicated exercises, like repeated practicing handwriting, to maintain some level of function. However, for some, these adaptions are insufficient, as activities once performed automatically, like climbing a ladder as on participant noted, are no longer within their physical capability.

Overall, these insights underscore the progressive nature of PD-related mobility challenges and highlight the need for targeted interventions to support patients in maintaining function and independence in daily life.

### Activities of Daily Living

The findings from the focus group discussion illustrate the significant impact of PD on activities of daily living, necessitating substantial lifestyle adjustments among patients. Fatigue, postural instability, and balance issues emerged as primary concerns, influencing how individuals structure their daily routines, navigate social environments, and engage in leisure activities. Participants described experiencing profound fatigue, often requiring scheduled rest periods throughout the day. One individual noted the tendency to nap around noon, highlighting how energy conservation becomes a key component of managing daily tasks. The need for increased planning and time management was also emphasized, with one participant explaining that completing routine activities, such as shopping or getting ready for an outing, now required significantly more time. Rather than approaching tasks spontaneously, individuals found it necessary to plan well in advance to accommodate the slower pace imposed by their condition.

In addition to modifying daily schedules, participants described how PD had led them to abandon previously enjoyed hobbies, particularly sports. Balance impairments were frequently cited as the main barrier to continued participation in activities such as hockey, baseball, and golf. One participant explicitly stated that they had to give up playing hockey in their leisure due to balance difficulties, while another noted that a variety of sports had become inaccessible as a result of postural instability. The loss of these activities was framed as inevitable consequences of disease progression, reflecting the broader impact of PD on quality of life and personal identify.

Participants also described modifications to how they engage with public spaces. For example, one individual reported choosing shorter checkout lines or avoiding crowded areas when grocery shopping, opting for spaces that allow for greater physical stability and ease of movement. This behavioral adaptation underscores how PD influences not only mobility but also social interactions and engagement with the environment. The decision to navigate spaces differently reflects an awareness of potential mobility challenges and the proactive strategies individuals develop to maintain independence.

A recurring theme in the discussion was the frustration associated with the deterioration of handwriting, which some participants found particularly distressing. While daily activities may remain largely unchanged for some, the inability to write legibly was perceived as a significant loss, affecting both personal expression and practical communication. The distress associated with losing the ability to write legibly is consistent with literature on the psychosocial burden of PD, where the loss of fine motor control can affect self-esteem, communication, and social interactions.^77^

Overall, these findings highlight the far-reaching consequences of PD on daily life, emphasizing the interplay between physical symptoms and behavioral adaptations. The loss of previously routine activities, whether recreational or function, underscores the progressive nature of PD and the continuous need for adjustments to maintain autonomy. The accounts provided by participants suggest that while individuals with PD develop strategies to cope with their symptoms, the impact on their social and physical engagement remains substantial. These insights reinforce the importance of supportive interventions, and structured fatigue management programs to help individual navigate the evolving challenges associated with PD.

### Emotional Wellbeing

Attitudes towards their diagnoses and pharmacological interventions also varied. Some participants demonstrated a determined, positive outlook, and others expressed resignation and frustration as the disease progresses. Some viewed medications, such as Levodopa, as life-changing, restoring a sense of normalcy, while others grew frustrated with the need for frequent adjustments or increased doses. Overall, some participants demonstrated more resilience and had a more positive outlook towards their disease progression than others. Furthermore, participants expressed that uncertainty about their future often leads to anxiety and worry, which are common feelings among PD patients.^44–47^ Ultimately, these symptoms can contribute to lower self-esteem, reduced self-efficacy, and a negative perception of one’s future health, causing many patients to develop a pessimistic outlook towards their disease progression.^48–50^ Integrating specialized mental healthcare by allying GPs with mental health clinicians can reduce the impact of these symptoms on the QoL of people with PD.^51^ This underscores the importance of addressing these symptoms throughout a patient’s treatment plan to improve long-term outcomes.

### Stigma

The experiences shared by participants highlight the impact of perceived and experienced stigma associated with PD. Participants expressed reluctance to disclose their diagnosis, fearing negative perceptions from others. One participant admitted to avoiding the use of assistive devices, such as canes, despite acknowledging their potential benefits. This hesitation was attributed to concerns about how they would be perceived, demonstrating the role of self-stigma in shaping behaviors and decisions regarding disease management. Similarly, another participant described being hesitant to inform others about their diagnosis, indicating a broader concern about how revealing their condition might affect social interactions or perceptions of their capabilities.

Beyond personal experiences, participants also recounted instances of observed stigma in their communities. One individual described witnessing a person with mobility challenges being mocked for using ski poles, which were likely needed for support. The participant felt compelled to advocate for the individual, however, their efforts were met with resistance. This reflects a possible broader societal lack of awareness and understanding regarding the use of adaptive aids.

These findings emphasize that stigma, both perceived and experienced, plays a role in the lives of individuals with PD. The fear of judgement can deter people for openly discussing their diagnosis or utilizing necessary assistive devices, potentially limiting their access to support and resources. The reluctance to disclose PD may also stem from concerns about being treated differently or facing assumptions abo9ut their abilities. Given that stigma was a prominent topic throughout the focus group, these insights underscore the need for greater public education and advocacy efforts to normalize PD symptoms and assistive devices, reducing stigma and fostering a more inclusive environment for those living with the disease.

### Social Support

Participants shared various experiences with caregivers, or lack thereof, as well as other forms of social support. Caregivers served an important role in most participants’ lives, from supporting participants during appointments, or reminding them to do things. In addition, participants elaborated on their supportive friendships as well as participation in community events such as walks for a cause. This emphasis on the importance of social and community support in reducing feelings of isolation and improving QoL is similarly supported in recent literature.^14, 59–62^ For instance, a cross-sectional study by Artigas et al. (2015) found that PD patients who attended peer support groups had significantly higher QoL and lower depression and anxiety scores compared to those who did not.^63^ Such support groups are particularly impactful for PD patients living in remote areas, as limited access to healthcare resources makes these groups a vital source of information along with improving the psychosocial wellbeing of patients.^64^ Establishing PD-specific social networks can not only enhance QoL outcomes, but can also help mitigate stigma by normalizing the experience and increasing public awareness of PD.^65^

### Other Related Symptoms

Cognitive challenges, such as memory deficits that require external assistance to manage daily tasks, alongside mental health concerns like anxiety about the uncertain progressions of PD were described by participants. Comorbid conditions such as Alzheimer’s Disease, sleep apnea, and arthritis that further complicate symptom management for some participants, highlighting the unique complicities of coexisting diagnoses. PD also manifests through a range of other symptoms, that can significantly impact lifestyle. For instance, participants described challenges such as changes in their voice and loss of smell. Despite hurdles in bodily discomfort, some participants describe the importance of exercise as an effective strategy to improve physical well-being and maintain a sense of control over their symptoms.

Participants described how NMS such as fatigue, cognitive deficits, and mental health challenges made daily activities more difficult, such as remembering to take medications regularly or sacrificing hobbies like going on long walks. Such changes to daily life have been found to further diminish already reduced QoL.^2^ Despite these concerns, however, NMS can often go unnoticed or unaddressed by healthcare providers.^42, 43^ Healthcare professionals must comprehensively address these symptoms to mitigate their negative impact on PD patients’ overall well-being.

Another insight gained from participants was the complex role that co-morbidities play when living with PD. Participants reported that co-morbid conditions such as arthritis exacerbated motor deficits, while sleep apnea negatively affected energy levels and motor function. PD patients often live with various co-morbidities, including dementia, sleep disorders, and psychiatric disorders, which can further diminish QoL.^66–69^ The complex interactions between PD symptoms and those of other co-morbid conditions further emphasizes the need for a patient-centered, integrated approach to symptom management. This approach should involve interdisciplinary teams of healthcare specialists working together to develop treatment plans that offer a holistic understanding of each patient’s condition.^70^ Personalizing PD treatment plans for common co-morbidities could optimize patient outcomes by addressing the full spectrum of symptoms.^71–74^

### Healthcare Navigation

Participant’s difficulties navigating the healthcare system was a prevalent theme throughout the focus group study, particularly in accessing healthcare due to long wait times for appointments, delays in receiving a diagnosis, and geographical barriers. Some shared their experiences of advocating for themselves or their care receivers, particularly when initial consultations felt inadequate or dismissive. For example, one participant sought a second opinion after being prescribed medication without further discussion. Other emphasized the importance of informed decision-making, with some opting for alternative treatments like supplements over medication, guided by supportive neurologists. These experiences highlight the struggles and persistence required to navigate PD care effectively. Geographic barriers were a common concern, as most participants reported long travel times to see their neurologists, with one even utilizing private funding for accessible transportation to another city. These concerns echo findings from O’Shea et al. (2023) highlighting long travel times as a major barrier for PD patients seeking healthcare.^33^ In our study, long wait times were also reported, with participants typically waiting between six months to a year for specialist appointments. These delays hinder timely access to care, causing distress and potentially exacerbated symptoms.^21, 34, 35^ Diagnostic and treatment delays can subsequently delay interventions that may slow disease progression, negatively impacting their QoL.^36–39^ In order for the Ontario healthcare system to address these ongoing accessibility issues, especially in underserved areas, the healthcare burden must be reduced by training more specialists willing to practice in such areas.^40, 41^ Disjointed coordination between healthcare professionals, as reported by focus group participants and previous research, can further widen gaps in access to care. ^16, 37, 41^ The insights gained highlight the increasing need for collaboration, communication, and availability of healthcare services to meet the needs of this vulnerable population and improve their QoL outcomes.

Participants also shared varied experiences regarding their ability to make decisions and advocate for themselves. Patients’ involvement in their own care, in addition to their ability to advocate for their needs, can significantly influence perceived control and QoL outcomes.^52, 53^ Patient-centered care (PCC) involves actively engaging patients in their care, and has been associated with increased treatment adherence and more positive health outcomes.^53^ Another effective way to implement PCC is through shared decision-making (SDM) in treatment-planning, which involves providing patients clear, simple information and engaging patients in discussions until a consensus is reached. Challenges in implementing SDM and PCC in Canada include the decentralized healthcare system, leading to variations in the scope of practice across provinces and differences in treatment options covered by universal healthcare.^54^ Increasing funding for research on SDM and PCC can help address these challenges and ensure their inclusion in future guidelines for health professionals.^54^ Additionally, research assessing patient involvement and QoL in PD patients have found that poor health literacy can hinder patient autonomy and self-advocacy.^16, 37, 55^ Thus, patient education programs aimed at improving understanding of the disease should be incorporated into PD treatment plans, and have been supported by previous literature.^56, 57^ Ultimately, equipping patients with the knowledge and skills to manage their health challenges can enhance their feelings of autonomy and involvement.^53, 58^

### Limitations and Future Directions

This study has some limitations which must be considered when interpreting the results.

Given the restraints in time and resources, a single-session focus group was the most feasible approach to gather meaningful insights within the scope of the study. While multiple focus groups could have provided broader perspectives, the initial session allowed for an in-depth discussion of key challenges faced by those with PD in Southwestern Ontario. However, the small sample size (N=6) significantly limited the generalizability of our findings. Notably, the small sample size may have failed to capture the full range of PD patients’ experiences, thus limiting the impact of this work. The current findings serve as a foundation for future research which could expand on these insights with additional focus groups or complimentary methodologies. Of note, because Woodstock, Ontario falls on the border of being an urban and rural community, with a population of approximately 50,000 and minimum of an hour drive to a major city, it is unclear whether these findings are unique to this population or other remote populations in Southwestern Ontario.^23^ Further exploratory research should focus on such regions, as they may present both unique and similar challenges to PD participants living in rural and urban communities.

Due to resource and time restraints, data saturation was not a primary objective of this study. While recurring patterns suggest that key themes were well-developed, future research with a larger or more diverse sample could further examine the saturation of these themes.

Moreover, future research could benefit from a mixed-methods approach to allow for correlations between quantitative factors such as age, time since diagnosis, symptom severity, and other demographics, and their relative impact on QoL.

In addition, the restraints in time and resources of the pilot study required physical attendance of participants which prevented individuals with mobility issues or lack of caregiver support to contribute to the focus group discussions. Given that mobility challenges were a key theme identified in this study, this exclusion may have unintentionally overlooked a crucial demographic. While resource limitations prevented a hybrid model in this pilot study, future research should explore virtual or hybrid formats to capture a wide range of individuals that can speak on their experiences living with PD. Additionally, the study sample included only one female participant, which limited the ability to compare male and female experiences. Previous literature has shown that PD impacts the QoL in men and women differently.^75^ Addressing gender disparities in future studies will be crucial for a more comprehensive understanding of PD’s impact on QoL.

Finally, this study’s recruitment method relied exclusively on PSSO members. While this convenience sampling was the most feasible approach given the time and resource constraints of this pilot study, it may have introduced bias and confounding variables. Participants who are PSSO members likely use the organization as a form of social support, which could positively influence their perceived QoL. Social support, as highlighted by the PDQ-8, plays a critical role in improving QoL.^76^ As such, the experiences of individuals who lack access to similar resources or support systems may have not been fully captured in this pilot study. Future studies should aim to include participants beyond formal support group networks to better represent the broader PD population. While a qualitative focus group provided valuable exploratory insights, future research could benefit from a mixed-methods approach. Combining qualitative data with quantitative measures—such as age, time since diagnosis, severity of symptoms, and other demographics—would allow for deeper analyses of the factors influencing QoL in individuals with PD.

## Conclusion

This study investigated the impact of PD on QoL through an exploratory focus group session. These findings provided unique insights from a specific population in Southwestern Ontario that has yet to be consulted in the literature. This study also took place in a familiar setting where PSSO focus groups were routinely held, which allowed participants to share freely in an environment where they felt supported by one another. The study procedures were exploratory in nature, which allowed the researchers to interact directly with the population of interest to obtain insights that may not have been captured using validated surveys such as the PDQ-8 or PDQ-39. Despite its limitations, this study provided preliminary insights into QoL experiences for individuals with PD and highlights important directions for future research.

Conducting this exploratory work, even with limited resources, allowed us to gather meaningful perspectives that can serve as a foundation for future research aimed at better understanding the QoL of PD populations living in Southwestern Ontario. These findings underscore the importance of a holistic, patient-centered approach to PD care. Interdisciplinary healthcare teams should collaborate to address all symptoms, including those related to PD and those associated with co-morbidities. It is also crucial to prioritize the mental and physical wellbeing of patients throughout their treatment plans. Addressing disparities in healthcare access and expanding social support networks will be vital in optimizing patient outcomes. A comprehensive, multidisciplinary approach is crucial for enhancing the QoL of PD patients.

## Supporting information

Focus Group Questions

Demographic Questionnaire

## Data Availability

All data produced in the present study are available upon reasonable request to the authors.

