## Supplementary material for "Quality of Life in Parkinson’s Disease: Insights from a Single-Session Focus Group in Southwestern Ontario": Focus Group Questions

**S1 Focus Group Questions**

1. Think about performing your daily activities. Can you recall any activities that you once did in your ‘spare’ or ‘leisure’ time, but now find hard to do?
2. Can you describe your experience with pain, and what therapies, activities, or interventions that you find to be the most effective in managing any of the painful symptoms you might have?
3. Have you experienced general feelings of anxiety, worry, feeling down or low towards visiting your doctor or specialist, hearing back from different healthcare professionals, or booking appointments with support service providers?
4. Do you ever find that you have a hard time remembering things, or concentrating during appointments with a healthcare provider?
5. For those of you who have a caregiver, how has the presence of this person impacted your everyday activities? For those of you who do not, in what aspect of your daily life would benefit most from having support?
6. Have you ever felt that you had to conceal your Parkinson's from people? If so, would you be open to sharing some of these experiences?
7. Have you ever experienced difficulties in being able to communicate your experiences to your healthcare provider, or felt that your concerns were being ignored due to these challenges?
8. Can you describe some of challenges you may have experienced with your mobility? (If examples are needed: moving around from place to place, maintaining your balance, fine motor movements of your hands, etc.).
9. Think back to when you received your diagnosis of PD, tell us about your experience in receiving this diagnosis. Was there anything that may have hindered or facilitated this process?
10. Out of all the things we discussed today, what factor impacting your everyday life do you feel is the most significant?
11. Out of all the things we discussed today, is there anything that we may have missed or anything that you may want to add to the discussion that was not talked about?
