## Supplementary material for "Quality of Life in Parkinson’s Disease: Insights from a Single-Session Focus Group in Southwestern Ontario": Demographic Questionnaire

**S3 Demographic Questionnaire**

| ANSWER THE FOLLOWING QUESTIONS AS THEY RELATE TO THE **PATIENT** |
|---|
| 1. When were you diagnosed with PD? ___________________________ |
| 2. How long have the symptoms of PD impacted your life? ___________________________ |
| 3. What is your age? ___________________________ |
| 4. What is your biological sex? ___________________________ |
| 5. What is your gender (identity)? ___________________________ |
| 6. What is your race? ___________________________ |
| 7. What is your ethnicity? ___________________________ |
| 8. What is your work status? ☐ Retired ☐ Working Part-Time ☐ Working Full-Time ☐ Other __________ |
| 9. What is your marital status? ☐ Single ☐ Married ☐ Widower ☐ Divorced ☐ Other ____________ |
| 10. What is your highest level of education? ☐ High School ☐ College ☐ University ☐ Other ______________ |
| 11. Has a doctor diagnosed you with any other chronic health conditions? If so, please list all that apply: ____________________________________________________________________________ |

**DO NOT FILL OUT (FOR RESEARCHER'S PURPOSES ONLY)**: Participant ID ___________________
